# Transient Apical Sparing in Hypertensive Heart Disease Explained by Laplace’s Law

**DOI:** 10.64898/2026.07.16.26358114

**Authors:** In-Chang Hwang, Hyue Mee Kim, Yeonggul Jang, Minjung Bak, Jiesuck Park, Jaeik Jeon, Seung-Ah Lee, Hong-Mi Choi, Yeonyee E. Yoon, Goo-Yeong Cho

## Abstract

**Background:** Apical sparing of left ventricular longitudinal strain (LS) is an echocardiographic clue to cardiac amyloidosis but may also occur in hypertensive heart disease (HHD).

**Objectives:** To determine whether apical sparing in HHD is associated with regional left ventricular wall stress estimated according to Laplace’s law.

**Methods:** We retrospectively studied 1,559 patients with HHD, 47 with light-chain cardiac amyloidosis (ALCA), and 409 normotensive controls. Artificial intelligence–assisted echocardiography quantified segmental LS, wall thickness, and cavity radius at the basal, midventricular, and apical levels. Wall stress was estimated as mean blood pressure (MBP) × radius/(2 × wall thickness). Apical sparing was defined as a relative regional strain ratio (RRSR)≥1.0.

**Results:** Apical sparing was present in 14 patients with HHD (0.9%), 13 with ALCA (27.7%), and no controls. Among HHD patients with apical sparing, RRSR decreased from 1.11±0.13 to 0.72±0.10 after antihypertensive treatment (*P*<0.001), accompanied by reduced wall stress and improved basal and midventricular LS, with resolution of apical sparing in all 14 patients. In the overall HHD cohort, changes in MBP and left ventricular mass index were independently associated with changes in RRSR. In an exploratory analysis of HHD patients with apical sparing, a reduction in basal wall stress was associated with a reduction in RRSR (β=0.267 for △RRSR×100, 95% CI 0.023–0.511; *P*=0.036). In ALCA, favorable hematologic response was the only determinant of RRSR reduction.

**Conclusions:** Apical sparing in HHD was uncommon but reversible and may represent a load-sensitive deformation pattern associated with regional wall stress, consistent with Laplace’s law.

**Condensed Abstract:** Apical sparing of left ventricular longitudinal strain occurred in 0.9% of patients with hypertensive heart disease and resolved in all affected patients after antihypertensive treatment. Resolution was accompanied by reduced regional wall stress and improved basal and midventricular strain, and reduction in basal wall stress was associated with reduction in the relative regional strain ratio. In contrast, apical sparing was more prevalent in light-chain cardiac amyloidosis and was related to disease stage and hematologic response. These findings support Laplace’s law as a physiologic framework for understanding reversible, load-sensitive apical sparing in hypertensive heart disease.

## Introduction

Apical sparing of left ventricular (LV) longitudinal strain (LS) on speckle-tracking echocardiography is recognized as a characteristic feature of cardiac amyloidosis and a red-flag sign necessitating downstream tests.(1) Recent ASE/EACVI clinical consensus also describes apical sparing of regional LS as a finding suggestive of possible cardiac amyloidosis.(2) Several mechanisms have been proposed, including lower amyloid burden at the apex, regional differences in myocardial fiber and matrix orientation, and greater basal vulnerability to remodeling related to turbulent flow and higher parietal stress.(3,4)

However, apical sparing is not pathognomonic for cardiac amyloidosis. Previous studies have reported this pattern in patients without cardiac amyloidosis, including those with severe aortic stenosis, chronic kidney disease, and hypertensive emergency.(5–8) These findings suggest that apical sparing should be interpreted as a deformation phenotype rather than a disease-specific diagnosis. Further, its occurrence in hypertension raises the possibility that pressure overload and LV geometry contribute to this strain pattern.

We hypothesized that Laplace’s law may provide a mechanistic explanation for apical sparing in selected patients with hypertensive heart disease (HHD). According to Laplace’s law, LV wall stress is proportional to pressure and cavity radius and inversely proportional to wall thickness. Because the LV radius is generally larger at the basal and midventricular levels than at the apex, pressure overload may impose greater wall stress and more impaired LS in these regions, resulting in relative apical preservation.(9,10) To test this hypothesis, we used artificial intelligence–assisted echocardiographic analysis to quantify segmental LV wall thickness, cavity radius, wall stress, and LS.(11) Using retrospective cohorts of HHD and AL cardiac amyloidosis (ALCA), with normotensive individuals with normal LV geometry as controls, we investigated the prevalence, reversibility, and wall-stress–based mechanism of apical sparing in HHD.(12–16)

## Methods

### Study population

We included three study groups: patients with hypertensive heart disease, patients with light-chain cardiac amyloidosis, and normotensive normal controls. The study was conducted in accordance with the Declaration of Helsinki and approved by the institutional review boards of the participating centers. The requirement for informed consent was waived due to the retrospective nature of the study and minimal risk to participants.

The hypertensive heart disease cohort was derived from the Strain for Risk Assessment and Therapeutic Strategies in Patients With Hypertensive Heart Disease (STRATS-HHD) Registry.(12–15) Consecutive patients with hypertension who underwent baseline echocardiography at Seoul National University Bundang Hospital or Chung-Ang University Hospital between 2006 and 2021 and follow-up echocardiography 6–18 months after antihypertensive treatment were screened. Patients with specific cardiomyopathies, ≥moderate valvular heart disease, end-stage renal disease, prior open-heart surgery, or cardiovascular diseases other than essential hypertension that could cause LV hypertrophy were excluded.(13) Of 1,872 registry participants, 1,559 with available baseline and follow-up segmental LS, wall thickness, radius, and estimated wall stress were included.

Patients with ALCA were used as a positive disease-control group and were derived from a retrospective cohort.(16) ALCA was diagnosed using serum and urine electrophoresis, serum free light-chain assay, cardiac magnetic resonance imaging, and tissue biopsy when clinically indicated.(17) Cardiac involvement was defined as mean LV wall thickness ≥12 mm without LV dilatation, accompanied by typical cardiac magnetic resonance findings. Patients with follow-up echocardiography 6–18 months after diagnosis were included. Treatment included chemotherapy and/or autologous stem cell transplantation at the attending physician’s discretion, and hematologic response was classified as complete remission, very good partial response, or no response according to established criteria.(18,19) In total, 47 patients with ALCA were included.

Normotensive individuals with normal LV geometry were identified from a previously established echocardiographic cohort and served as normal controls.(20) Because follow-up echocardiography was unavailable, only baseline data were analyzed. A total of 409 controls were included.

### Echocardiography

Comprehensive two-dimensional, M-mode, and Doppler echocardiography was performed using commercially available ultrasound systems. Conventional measurements were obtained according to American Society of Echocardiography guidelines.(21) LV mass was calculated using the Devereux formula, and LV mass index (LVMI) was derived by normalizing LV mass to body surface area, with LV hypertrophy (LVH) defined as LVMI >115 g/m² in men or >95 g/m² in women.(22) Relative wall thickness (RWT) of >0.42 indicated concentric geometry. LV geometry was classified as normal, concentric remodeling, concentric hypertrophy, or eccentric hypertrophy according to LVMI and RWT. LV ejection fraction (LVEF) was calculated by Simpson’s biplane method, and LA volume index (LAVI) was obtained by dividing LA volume by body surface area.

LV and LA strain were quantified using an AI-based echocardiographic analysis system (Sonix Health Workstation, Version 2.0; Ontact Health Co., Ltd., Korea), which provides automated view classification, segmentation, motion tracking, and parameter extraction.(11,23–25) Specifically, automated LV strain analysis was performed using the Segmentation-based Myocardial Advanced Refinement Tracking (SMART) system, a fully automated deep learning–based method integrating myocardial segmentation and speckle-tracking–based motion estimation.(11) After frame-by-frame myocardial segmentation, motion vectors were iteratively refined by aligning the mid-myocardial line and correcting endocardial and epicardial borders throughout the cardiac cycle. LV global LS was calculated as the mean peak LS from apical 4-, 2-, and 3-chamber views, referenced to the QRS complex.(23) Detailed information on the development and validation of the SMART system has been published previously.(11)

The SMART-derived tracked contours were further used to automatically extract segmental LV geometry, including myocardial wall thickness and LV internal radius at the basal, midventricular, and apical levels from apical views without manual correction. Measurements were obtained on an 18-segment basis from apical 4-, 2-, and 3-chamber views and averaged at basal, midventricular, and apical levels (**Central illustration**). To estimate LV wall stress at the basal (*T_base_*), midventricular (*T_mid_*), and apical (*T_apex_*) levels, the average LV wall thickness at end-diastole and the average LV internal radius at end-diastole for each level were used. As a surrogate for LV pressure, mean LV pressure was assumed to be equivalent to mean blood pressure (MBP), calculated as MBP = (SBP + 2 × DBP) / 3, given that patients with significant valvular heart disease were excluded. Segmental LV wall stress (σ) at each level was then calculated according to Laplace’s law:

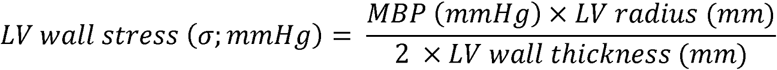

Apical sparing was quantified using the relative regional strain ratio (RRSR), and apical sparing was defined as RRSR ≥1.0:

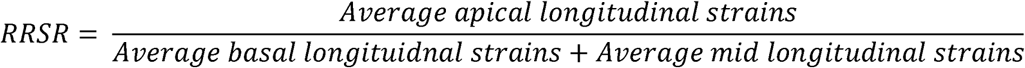

### Outcomes

The primary analyses evaluated the relationships among segmental LV geometry, estimated wall stress, and segmental LS in HHD, ALCA, and normal controls. The main outcomes were:

1) baseline prevalence of apical sparing; 2) temporal changes in wall stress, segmental LS, and RRSR; and 3) predictors of baseline apical sparing and change in RRSR.

### Statistical analysis

Continuous variables are presented as mean±SD or median with interquartile range, and categorical variables as numbers and percentages. Baseline characteristics were compared among groups using analysis of variance or the Kruskal–Wallis test for continuous variables and the chi-square test or Fisher exact test for categorical variables, as appropriate. Segmental wall thickness, LV radius, estimated wall stress, and segmental LS were compared across LV levels and etiologic groups. Because segmental measurements were clustered within patients, linear mixed-effects models with a patient-specific random intercept were used when appropriate. Temporal changes between baseline and follow-up echocardiography were assessed using paired statistical tests.

Predictors of baseline apical sparing were evaluated separately in the HHD and ALCA groups using Firth penalized logistic regression with parsimonious, clinically prespecified models. Predictors of change in RRSR were evaluated separately in the entire HHD and ALCA populations using multivariable linear regression with clinically prespecified covariates. Candidate variables were selected based on clinical relevance and the proposed pathophysiologic framework, and variables with substantial conceptual or mathematical overlap were not included simultaneously in the same model. Because of limited sample sizes, analyses restricted to patients with baseline apical sparing used exploratory univariable linear regression. Favorable hematologic response, defined as complete response or very good partial response, was included in analyses of the ALCA group. Multicollinearity was assessed before model construction.

For improved interpretability, ΔRRSR was multiplied by 100 before linear regression analysis; therefore, the regression coefficients represent the change in RRSR ×100. Adjusted odds ratios, regression coefficients, and 95% confidence intervals were reported. All analyses were performed using R software, and two-sided *P* values <0.05 were considered statistically significant.

## Results

### Baseline characteristics

The study population included 1,559 patients with HHD, 47 with ALCA, and 409 normal controls (**Table 1**). Baseline characteristics of three groups, stratified by the presence of apical sparing at baseline, are summarized in **Table 1**. Patients with ALCA were oldest, followed by those with HHD and normal controls (71.4±9.2, 65.0±13.2, and 48.2±11.8 years; P<0.001). The proportion of men was similar in HHD and ALCA but lower in normal controls (60.9%, 61.7%, and 42.3%; P<0.001). Systolic, diastolic, and mean BPs were highest in HHD and lowest in ALCA (all P<0.001).

**Table 1.** Baseline characteristics. Values are presented as mean ± SD or n (%), unless otherwise indicated. * *P* values compare patients with HHD without versus with apical sparing at baseline. † *P* values compare patients with ALCA without versus with apical sparing at baseline. ‡ *P* values compare the overall HHD, ALCA, and normal-control groups. Continuous variables were compared using analysis of variance or the Kruskal–Wallis test, as appropriate, and categorical variables using the chi-square test or Fisher exact test, as appropriate. § Renal impairment was defined as an estimated glomerular filtration rate <60 mL/min/1.73 m² at baseline. Apical sparing was defined as an RRSR ≥1.0. Treatment categories for ALCA were not mutually exclusive. N/A indicates that the variable or statistical comparison was not applicable. ALCA, light-chain cardiac amyloidosis; AutoSCT, autologous stem cell transplantation; CCB, calcium channel blocker; CR, complete response; DBP, diastolic blood pressure; HHD, hypertensive heart disease; IVSd, interventricular septal thickness at end-diastole; LA, left atrial; LAVI, left atrial volume index; LV, left ventricular; LV-EDD, left ventricular end-diastolic diameter; LV-EDV, left ventricular end-diastolic volume; LV-EF, left ventricular ejection fraction; LV-ESD, left ventricular end-systolic diameter; LV-ESV, left ventricular end-systolic volume; LV-GLS, left ventricular global longitudinal strain; LV-MI, left ventricular mass index; LVPWd, left ventricular posterior wall thickness at end-diastole; MBP, mean blood pressure; MRA, mineralocorticoid receptor antagonist; N/A, not applicable; PD, progressive disease; PI, proteasome inhibitor; PR, partial response; RAS, renin–angiotensin system; RRSR, relative regional strain ratio; RVSP, right ventricular systolic pressure; RWT, relative wall thickness; SBP, systolic blood pressure; SD, stable disease; STRATS-HHD, Strain for Risk Assessment and Therapeutic Strategies in Patients With Hypertensive Heart Disease; TR Vmax, peak tricuspid regurgitation velocity; VGPR, very good partial response.

| Variables | STRATS-HHD<br>(n = 1,559) |  |  |  | ALCA<br>(n=47) |  |  |  | Normal<br>(n=409) | P‡ |
| --- | --- | --- | --- | --- | --- | --- | --- | --- | --- | --- |
|  | Total | without | with | P* | Total | without | with | P† |  |  |
|  |  | apical sparing<br>(n = 1,545) | apical sparing<br>(n = 14) |  |  | apical sparing<br>(n = 34) | apical sparing<br>(n = 13) |  |  |  |
| Clinical Factors |  |  |  |  |  |  |  |  |  |  |
| Age (years) | 65.0 ± 13.2 | 65.2 ± 13.1 | 51.8 ± 14.6 | <0.001 | 71.4 ± 9.2 | 72.4 ± 9.4 | 68.6 ± 8.3 | 0.212 | 48.2 ± 11.8 | <0.001 |
| Male sex | 949 (60.9%) | 940 (60.8%) | 9 (64.3%) | 0.793 | 29 (61.7%) | 19 (55.9%) | 10 (76.9%) | 0.315 | 173 (42.3%) | <0.001 |
| Hypertension | 1,559 (100.0%) | 1,545 (100.0%) | 14 (100.0%) | N/A | 23 (48.9%) | 15 (44.1%) | 8 (61.5%) | 0.285 | 0 (0.0%) | <0.001 |
| Diabetes mellitus | 449 (28.8%) | 447 (28.9%) | 2 (14.3%) | 0.228 | 10 (21.3%) | 8 (23.5%) | 2 (15.4%) | 0.703 | 0 (0.0%) | <0.001 |
| Dyslipidemia | 442 (28.4%) | 439 (28.4%) | 3 (21.4%) | 0.844 | 16 (34.0%) | 11 (32.4%) | 5 (38.5%) | 0.693 | 0 (0.0%) | <0.001 |
| Renal impairment§ | 343 (22.0%) | 335 (21.7%) | 8 (57.1%) | 0.004 | 18 (38.3%) | 14 (41.2%) | 4 (30.8%) | 0.739 | 0 (0.0%) | <0.001 |
| Coronary artery disease | 592 (38.0%) | 589 (38.1%) | 3 (21.4%) | 0.200 | 1 (2.1%) | 1 (2.9%) | 0 (0.0%) | 0.723 | 0 (0.0%) | <0.001 |
| Atrial fibrillation | 208 (13.4%) | 208 (13.5%) | 0 (0.0%) | 0.140 | 8 (17.0%) | 3 (8.8%) | 5 (38.5%) | 0.028 | 0 (0.0%) | <0.001 |
| Stroke | 201 (12.9%) | 197 (12.8%) | 4 (28.6%) | 0.208 | 5 (10.6%) | 2 (5.9%) | 3 (23.1%) | 0.121 | 0 (0.0%) | <0.001 |
| Anthropometric Factors |  |  |  |  |  |  |  |  |  |  |
| Body mass index (kg/m²) | 25.1 ± 3.6 | 25.1 ± 3.6 | 23.9 ± 5.2 | 0.222 | 24.1 ± 2.5 | 23.6 ± 2.5 | 23.3 ± 2.4 | 0.717 | 23.5 ± 2.9 | <0.001 |
| SBP at baseline (mmHg) | 153.1 ± 24.4 | 152.7 ± 24.0 | 188.5 ± 34.2 | <0.001 | 113.7 ± 20.6 | 114.3 ± 19.3 | 112.3 ± 24.5 | 0.774 | 120.2 ± 10.5 | <0.001 |
| DBP at baseline (mmHg) | 89.7 ± 18.3 | 89.4 ± 17.9 | 120.6 ± 32.6 | <0.001 | 68.4 ± 9.8 | 68.4 ± 8.5 | 68.5 ± 13.0 | 0.962 | 72.6 ± 7.6 | <0.001 |
| MBP at baseline (mmHg) | 110.8 ± 18.9 | 110.5 ± 18.5 | 143.3 ± 32.4 | <0.001 | 83.5 ± 12.4 | 83.7 ± 10.7 | 83.1 ± 16.4 | 0.891 | 88.4 ± 7.9 | <0.001 |
| Baseline echocardiography |  |  |  |  |  |  |  |  |  |  |
| IVSd (mm) | 10.7 ± 2.1 | 10.7 ± 2.1 | 12.9 ± 2.4 | <0.001 | 14.3 ± 2.6 | 14.2 ± 2.8 | 14.6 ± 2.1 | 0.634 | 8.6 ± 1.2 | <0.001 |
| LV-EDD (mm) | 48.9 ± 6.3 | 48.9 ± 6.3 | 49.2 ± 8.4 | 0.832 | 42.8 ± 5.8 | 43.1 ± 6.2 | 42.0 ± 4.6 | 0.572 | 46.0 ± 3.8 | <0.001 |
| LV-ESD (mm) | 32.3 ± 7.7 | 32.3 ± 7.7 | 35.6 ± 9.1 | 0.107 | 30.8 ± 5.8 | 30.0 ± 6.1 | 33.1 ± 4.2 | 0.101 | 29.2 ± 3.7 | <0.001 |
| LVPWd (mm) | 10.3 ± 1.9 | 10.3 ± 1.9 | 13.3 ± 2.9 | <0.001 | 13.5 ± 2.2 | 13.3 ± 2.3 | 13.9 ± 2.1 | 0.363 | 8.3 ± 1.1 | <0.001 |
| LV-MI (g/m²) | 109.5 ± 33.6 | 109.1 ± 33.3 | 151.5 ± 46.8 | <0.001 | 138.9 ± 33.9 | 140.1 ± 37.2 | 135.7 ± 24.4 | 0.698 | 74.7 ± 13.4 | <0.001 |
| RWT | 0.43 ± 0.10 | 0.43 ± 0.09 | 0.54 ± 0.10 | <0.001 | 0.64 ± 0.15 | 0.63 ± 0.15 | 0.68 ± 0.16 | 0.336 | 0.36 ± 0.05 | <0.001 |
| LV-EDV (mL) | 85.4 ± 35.7 | 85.2 ± 35.6 | 111.5 ± 43.2 | 0.006 | 70.0 ± 24.2 | 70.5 ± 24.9 | 68.7 ± 23.0 | 0.828 | 80.4 ± 19.1 | 0.002 |
| LV-ESV (mL) | 38.4 ± 28.8 | 38.2 ± 28.8 | 57.0 ± 30.1 | 0.015 | 31.7 ± 15.5 | 31.8 ± 16.9 | 31.6 ± 11.5 | 0.977 | 29.1 ± 7.8 | <0.001 |
| LV-EF (%) | 58.7 ± 11.5 | 58.7 ± 11.4 | 51.0 ± 13.2 | 0.012 | 55.9 ± 8.9 | 56.7 ± 9.8 | 53.9 ± 5.8 | 0.347 | 63.9 ± 4.0 | <0.001 |
| LA diameter (mm) | 39.7 ± 7.2 | 39.7 ± 7.2 | 42.0 ± 8.7 | 0.258 | 42.6 ± 5.4 | 41.7 ± 5.7 | 44.8 ± 3.5 | 0.078 | 34.2 ± 4.2 | <0.001 |
| LAVI (mL/m²) | 39.8 ± 21.6 | 39.6 ± 21.6 | 52.0 ± 15.2 | 0.034 | 54.1 ± 18.8 | 55.0 ± 19.4 | 51.6 ± 17.5 | 0.585 | 29.2 ± 5.4 | <0.001 |
| Mitral E/e' ratio | 12.8 ± 6.5 | 12.8 ± 6.5 | 16.2 ± 8.1 | 0.050 | 24.2 ± 8.1 | 24.2 ± 8.3 | 24.1 ± 8.1 | 0.981 | 7.6 ± 1.7 | <0.001 |
| LV-GLS (%) | -14.7 ± 3.6 | -14.7 ± 3.6 | -11.3 ± 3.0 | <0.001 | -10.3 ± 2.8 | -10.6 ± 3.1 | -9.6 ± 1.7 | 0.158 | -19.0 ± 1.6 | <0.001 |
| TR Vmax (m/s) | 2.4 ± 0.5 | 2.4 ± 0.5 | 2.5 ± 0.5 | 0.385 | 2.6 ± 0.4 | 2.6 ± 0.4 | 2.8 ± 0.5 | 0.180 | 2.2 ± 0.3 | <0.001 |
| RVSP (mmHg) | 30.1 ± 9.4 | 30.1 ± 9.4 | 32.4 ± 11.8 | 0.395 | 34.4 ± 10.1 | 33.1 ± 8.9 | 37.4 ± 12.3 | 0.206 | 24.5 ± 4.5 | <0.001 |
| LV geometry at baseline |  |  |  | <0.001 |  |  |  | 0.641 |  | <0.001 |
| Normal | 536 (34.4%) | 535 (34.6%) | 1 (7.1%) |  | 1 (2.1%) | 1 (2.9%) | 0 (0.0%) |  | 409 (100.0%) |  |
| Concentric remodeling | 328 (21.0%) | 326 (21.1%) | 2 (14.3%) |  | 7 (14.9%) | 6 (17.6%) | 1 (7.7%) |  | 0 (0.0%) |  |
| Concentric hypertrophy | 365 (23.4%) | 355 (23.0%) | 10 (71.4%) |  | 38 (80.9%) | 26 (76.5%) | 12 (92.3%) |  | 0 (0.0%) |  |
| Eccentric hypertrophy | 330 (21.2%) | 329 (21.3%) | 1 (7.1%) |  | 1 (2.1%) | 1 (2.9%) | 0 (0.0%) |  | 0 (0.0%) |  |
| **Antihypertensive medication** |  |  |  |  |  |  |  |  |  |  |
| RAS blockers | 1049 (67.3%) | 1037 (67.1%) | 12 (85.7%) | 0.140 | 14 (29.8%) | 8 (23.5%) | 6 (46.2%) | 0.129 | 0 (0.0%) | <0.001 |
| Beta blockers | 571 (36.6%) | 563 (36.4%) | 8 (57.1%) | 0.161 | 10 (21.3%) | 5 (14.7%) | 5 (38.5%) | 0.112 | 0 (0.0%) | <0.001 |
| CCB | 788 (50.5%) | 775 (50.2%) | 13 (92.9%) | 0.001 | 10 (21.3%) | 7 (20.6%) | 3 (23.1%) | 0.852 | 0 (0.0%) | <0.001 |
| Thiazide | 257 (16.5%) | 252 (16.3%) | 5 (35.7%) | 0.052 | 1 (2.1%) | 0 (0.0%) | 1 (7.7%) | 0.277 | 0 (0.0%) | <0.001 |
| MRA | 112 (7.2%) | 109 (7.1%) | 3 (21.4%) | 0.038 | 19 (40.4%) | 12 (35.3%) | 7 (53.8%) | 0.246 | 0 (0.0%) | <0.001 |
| **Chemotherapy for ALCA** |  |  |  |  |  |  |  |  |  |  |
| AutoSCT | N/A | N/A | N/A | N/A | 10 (21.3%) | 6 (17.6%) | 4 (30.8%) | 0.325 | N/A | N/A |
| Daratumumab-containing regimen | N/A | N/A | N/A | N/A | 2 (4.3%) | 1 (2.9%) | 1 (7.7%) | 0.470 | N/A | N/A |
| PI-based regimen without daratumumab | N/A | N/A | N/A | N/A | 33 (70.2%) | 24 (70.6%) | 9 (69.2%) | 0.927 | N/A | N/A |
| Non-PI combination or monotherapy | N/A | N/A | N/A | N/A | 16 (34.0%) | 12 (35.3%) | 4 (30.8%) | 0.770 | N/A | N/A |
| Revised Mayo Stage (2012) | N/A | N/A | N/A | N/A |  |  |  | 0.104 | N/A | N/A |
| - Stage I | N/A | N/A | N/A | N/A | 0 (0.0%) | 0 (0.0%) | 0 (0.0%) |  | N/A | N/A |
| - Stage II | N/A | N/A | N/A | N/A | 4 (8.5%) | 4 (11.8%) | 0 (0.0%) |  | N/A | N/A |
| - Stage III | N/A | N/A | N/A | N/A | 18 (38.3%) | 15 (44.1%) | 3 (23.1%) |  | N/A | N/A |
| - Stage IV | N/A | N/A | N/A | N/A | 25 (53.2%) | 15 (44.1%) | 10 (76.9%) |  | N/A | N/A |
| Response | N/A | N/A | N/A | N/A |  |  |  | 0.474 | N/A | N/A |
| - Complete response (CR) | N/A | N/A | N/A | N/A | 11 (23.4%) | 6 (17.6%) | 5 (38.5%) |  | N/A | N/A |
| - Very good partial response (VGPR) | N/A | N/A | N/A | N/A | 9 (19.1%) | 6 (17.6%) | 3 (23.1%) |  | N/A | N/A |
| - No response | N/A | N/A | N/A | N/A | 27 (57.4%) | 22 (64.7%) | 5 (38.5%) |  | N/A | N/A |

Values are presented as mean ± SD or n (%), unless otherwise indicated.
§ Renal impairment was defined as an estimated glomerular filtration rate <60 mL/min/1.73 m<sup>2</sup> at baseline.

LVMI, LAVI, and mitral E/e′ were highest in ALCA, whereas LVEF and LV global LS were most impaired in ALCA, followed by HHD and normal controls. LV global LS was −10.3 ± 2.8%, −14.7 ± 3.6%, and −19.0 ± 1.6%, respectively (P<0.001). Concentric LVH was present in 80.9% of patients with ALCA, whereas all normal controls had normal LV geometry.

### Segmental LV Geometry, Wall Stress, and Longitudinal Strain

Segmental LV wall thickness, radius, estimated wall stress, and LS are shown in **Supplementary Figure S1**. Wall thickness was greatest in ALCA at the basal and midventricular levels, whereas estimated wall stress was highest in HHD across all LV levels. Basal, midventricular, and apical LS values were −14.0 ± 4.0%, −13.9 ± 4.0%, and −16.4 ± 4.5% in HHD; −7.3 ± 3.9%, −9.8 ± 3.0%, and −13.5 ± 3.4% in ALCA; and −15.9 ± 3.0%,−17.7 ± 2.8%, and −20.1 ± 3.3% in normal controls.

Associations between wall stress and segmental LS differed by etiology (**Figure 1A**). In HHD, higher wall stress was associated with more impaired LS at all levels (basal slope 0.727, midventricular slope 1.023, and apical slope 0.650; all P<0.001). Conversely, higher wall stress was associated with more preserved LS in ALCA at the basal and midventricular levels and in normal controls at the basal and midventricular levels. These relationships persisted after stratification by baseline apical sparing (**Figures 1B** and **1C**). In HHD, patients with apical sparing had greater basal and midventricular LS impairment, whereas apical LS was comparable between those with and without apical sparing.

**Figure 1.**
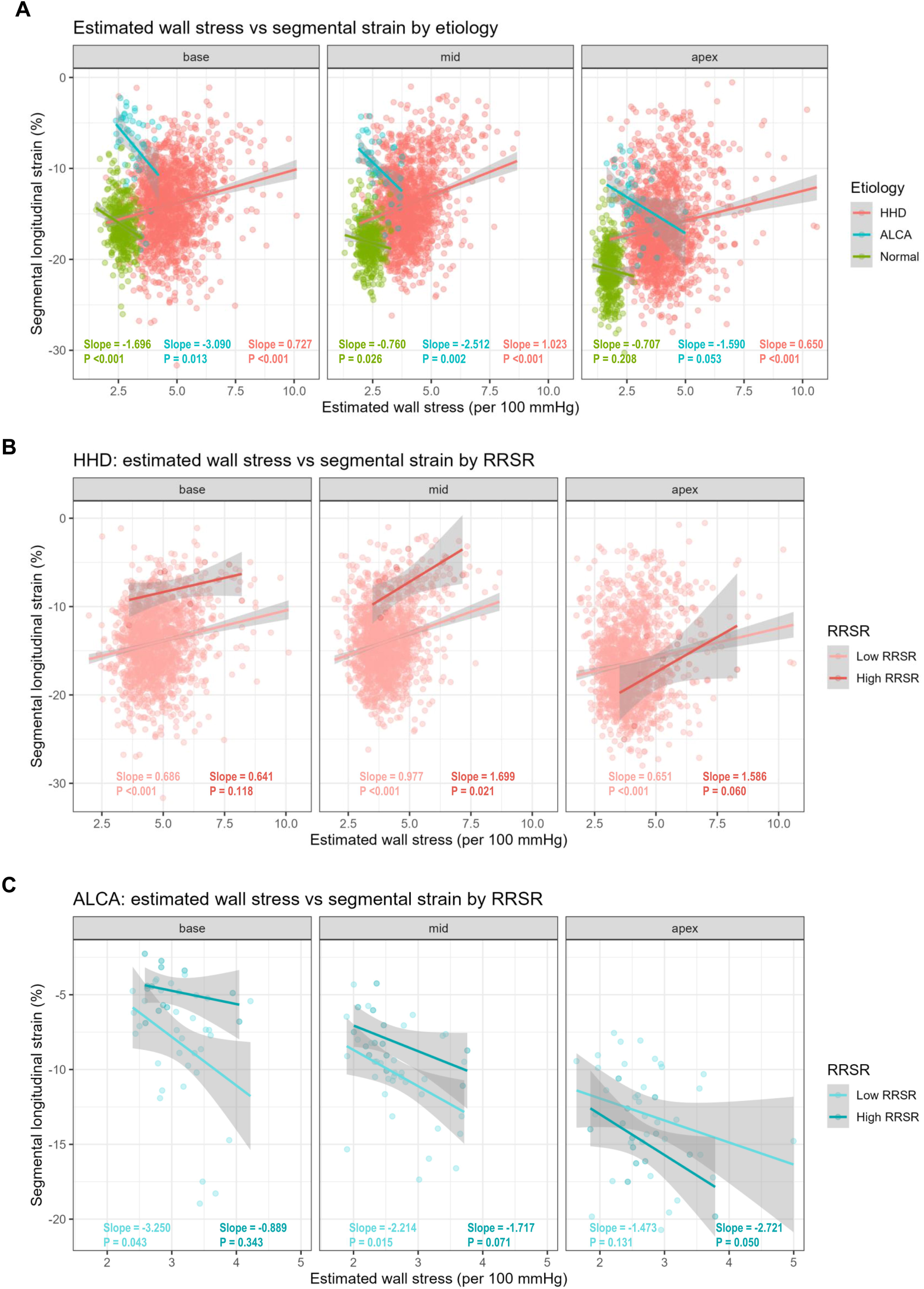
Associations of LV wall stress with segmental LS in HHD and ALCA. Associations at baseline are shown (A) across HHD, ALCA, and normal groups; (B) according to RRSR <1.0 versus ≥1.0 in HHD; and (C) according to RRSR <1.0 versus ≥1.0 in ALCA. Abbreviations: ALCA, light-chain cardiac amyloidosis; HHD, hypertensive heart disease; LS, longitudinal strain; LV, left ventricular; and RRSR, relative regional strain ratio.

### Prevalence and Temporal Changes of Apical Sparing

At baseline, mean RRSR was 0.60±0.15 in HHD, 0.85±0.25 in ALCA, and 0.62±0.09 in normal controls (**Figure 2A**). The prevalence of apical sparing decreased progressively as the RRSR cutoff increased from 0.7 to 1.0 in all groups (**Figure 2B**). From baseline to follow-up, mean RRSR and apical-sparing prevalence decreased in HHD but increased in ALCA. Using the conventional cutoff of RRSR ≥1.0, apical sparing was present at baseline in 14 of 1,559 patients with HHD (0.9%) and 13 of 47 with ALCA (27.7%), but in no normal controls. In HHD, patients with apical sparing were younger and had more renal impairment, higher BP, greater LVMI, and greater RWT than those without apical sparing (**Table 1**). No significant baseline differences were observed according to apical-sparing status in ALCA.

**Figure 2.**
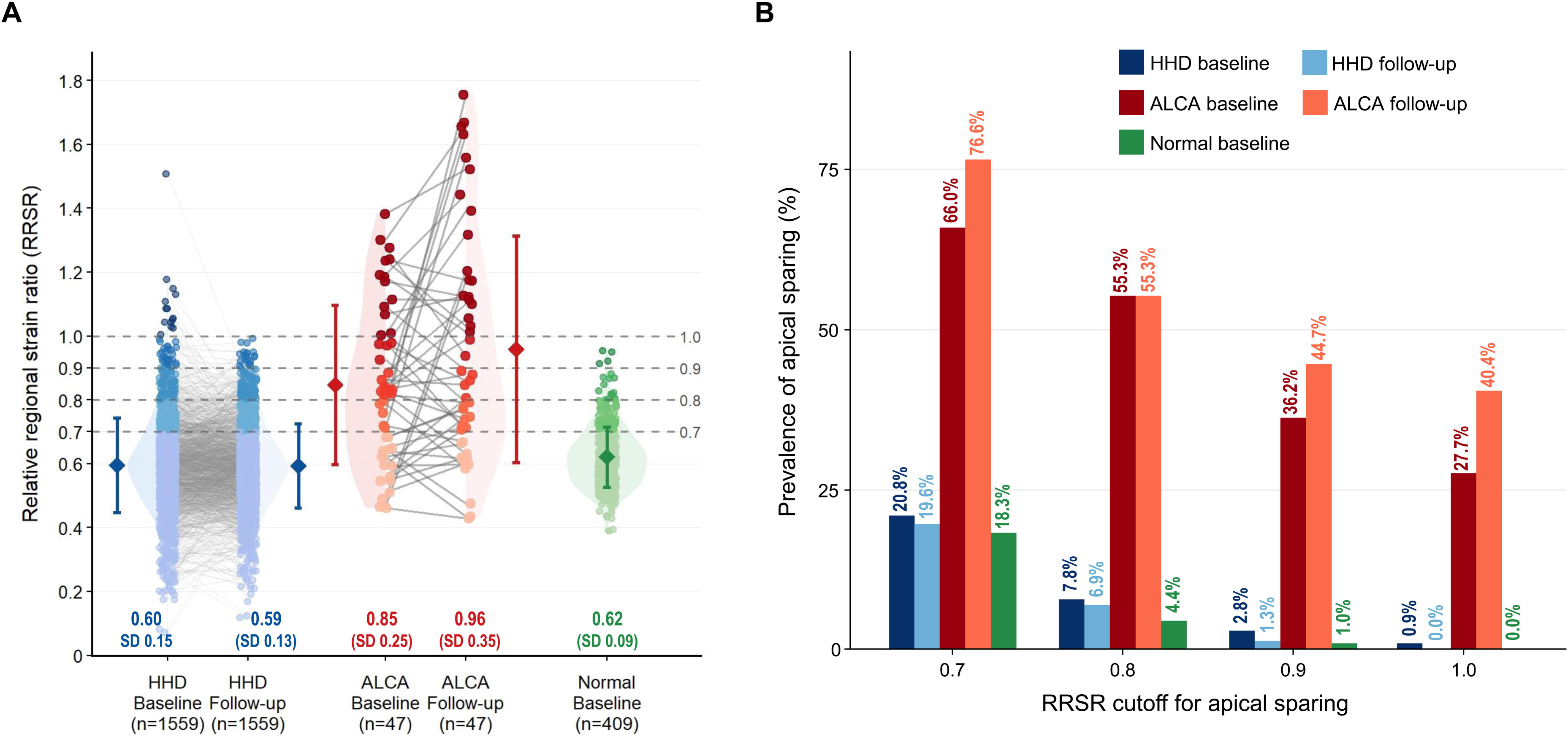
Temporal changes in RRSR and prevalence of apical sparing. (A) RRSR distributions at baseline and follow-up in HHD and ALCA and at baseline in normal controls. Grey lines connect paired measurements; diamonds and error bars indicate mean ± standard deviation. (B) Prevalence of apical sparing according to RRSR cutoffs of 0.7, 0.8, 0.9, and 1.0. Abbreviations as in Figure 1.

Predictors of baseline apical sparing are shown in Table 2. In HHD, younger age (adjusted OR: 0.619 per 10 years; 95% CI: 0.403–0.951; P=0.029), lower BMI (adjusted OR: 0.408 per 10 kg/m²; 95% CI: 0.198–0.841; P=0.015), higher MBP (adjusted OR: 1.373 per 10 mmHg; 95% CI: 1.110–1.698; P=0.003), and eGFR <60 mL/min/1.73 m² (adjusted OR: 5.218; 95% CI: 1.583–17.196; P=0.007) were independently associated with apical sparing. In ALCA, the 2012 revised Mayo stage was the only independent determinant (adjusted OR: 5.534 per stage; 95% CI: 1.249–24.527; P=0.024).

**Table 2.** Predictors of apical sparing pattern at baseline in HHD. Results are derived from Firth penalized logistic regression analyses using parsimonious, clinically prespecified models. Adjusted ORs are presented with 95% CIs. Given the limited number of patients with apical sparing, these analyses should be considered exploratory. Abbreviations: ALCA, light-chain cardiac amyloidosis; BMI, body mass index; CI, confidence interval; eGFR, estimated glomerular filtration rate; HHD, hypertensive heart disease; MBP, mean blood pressure; OR, odds ratio.

| HHD |  |  |  | ALCA |  |  |  |
| --- | --- | --- | --- | --- | --- | --- | --- |
| Variables | Adj. OR | 95 % CI | P-value | Variables | Adj. OR | 95% CI | P-value |
| Age (per +10 year) | 0.619 | 0.403 – 0.951 | 0.029 | 2012 Mayo stage (per +1 stage) | 5.534 | 1.249 – 24.527 | 0.024 |
| MBP at baseline (per +10 mmHg) | 1.373 | 1.110 – 1.698 | 0.003 |  |  |  |  |
| BMI at baseline (per +10 kg/m<sup>2</sup>) | 0.408 | 0.198 – 0.841 | 0.015 |  |  |  |  |
| eGFR <60 mL/min/1.73m<sup>2</sup> | 5.218 | 1.583 – 17.196 | 0.007 |  |  |  |  |

Detailed temporal changes according to baseline apical-sparing status are shown in **Tables 3 and 4**. In HHD without baseline apical sparing, segmental LS and wall stress improved modestly, whereas RRSR did not change significantly. Among the 14 patients with baseline apical sparing, RRSR decreased from 1.11±0.13 to 0.72±0.10 (P<0.001), with resolution of apical sparing in all patients. Wall stress decreased at all LV levels, and basal and midventricular LS improved significantly, whereas apical LS did not change significantly. In ALCA without baseline apical sparing, RRSR increased from 0.72 ± 0.15 to 0.88 ± 0.36 (P=0.012). RRSR remained unchanged among those with baseline apical sparing (1.18 ± 0.11 vs 1.17 ± 0.26; P=0.945). Wall stress decreased across all LV levels in patients without baseline apical sparing but remained largely unchanged in those with apical sparing.

**Table 3.** Temporal changes in echocardiographic parameters according to apical sparing pattern in patients with HHD. Values are presented as mean ± SD or n (%). Differences were calculated as follow-up minus baseline values. *P* values represent within-group comparisons between baseline and follow-up measurements using paired statistical tests. Apical sparing was defined as an RRSR ≥1.0 at baseline. Segmental LV radius, wall thickness, longitudinal strain, and estimated wall stress represent values averaged across the basal, midventricular, or apical segments from the apical 4-, 2-, and 3-chamber views. Segmental wall stress was estimated according to Laplace’s law as MBP × radius/(2 × wall thickness). Negative changes in longitudinal strain indicate that strain became more negative, corresponding to improved systolic deformation. Available paired observations were used for each variable; consequently, the mean within-patient difference may not equal the arithmetic difference between the separately reported baseline and follow-up means. Abbreviations: HHD, hypertensive heart disease; LA, left atrial; LAVI, left atrial volume index; LV, left ventricular; LV-EDD, left ventricular end-diastolic diameter; LV-EDV, left ventricular end-diastolic volume; LV-EF, left ventricular ejection fraction; LV-ESD, left ventricular end-systolic diameter; LV-ESV, left ventricular end-systolic volume; LV-GLS, left ventricular global longitudinal strain; LV-MI, left ventricular mass index; MBP, mean blood pressure; RRSR, relative regional strain ratio; RVSP, right ventricular systolic pressure; RWT, relative wall thickness; TR Vmax, peak tricuspid regurgitation velocity; σ, estimated wall stress.

| Variables | No apical sparing at baseline (n = 1,545) |  |  |  | Apical sparing (+) at baseline (n = 14) |  |  |  |
| --- | --- | --- | --- | --- | --- | --- | --- | --- |
|  | Baseline | Follow-up | Difference | P-value | Baseline | Follow-up | Difference | P-value |
| **Echocardiography** |  |  |  |  |  |  |  |  |
| LV-EDD (mm) | 48.9 ± 6.3 | 48.3 ± 5.7 | -0.6 ± 4.8 | <0.001 | 49.2 ± 8.4 | 45.2 ± 7.5 | -4.0 ± 5.8 | 0.022 |
| LV-ESD (mm) | 32.3 ± 7.7 | 31.1 ± 6.6 | -1.1 ± 5.8 | <0.001 | 35.6 ± 9.1 | 27.6 ± 7.9 | -8.0 ± 8.6 | 0.004 |
| LV-EDV (mL) | 85.2 ± 35.6 | 81.6 ± 34.9 | -3.7 ± 29.4 | <0.001 | 111.5 ± 43.2 | 88.8 ± 23.8 | -22.8 ± 36.6 | 0.037 |
| LV-ESV (mL) | 38.2 ± 28.8 | 34.1 ± 22.2 | -4.1 ± 19.7 | <0.001 | 57.0 ± 30.1 | 30.1 ± 7.5 | -26.9 ± 27.7 | 0.003 |
| LV-EF (%) | 58.7 ± 11.4 | 60.3 ± 9.6 | 1.6 ± 9.1 | <0.001 | 51.0 ± 13.2 | 65.9 ± 3.8 | 14.9 ± 11.7 | <0.001 |
| LV-MI (g/m<sup>2</sup>) | 109.1 ± 33.3 | 108.4 ± 32.7 | -0.8 ± 33.2 | 0.346 | 151.5 ± 46.8 | 106.9 ± 30.8 | -44.6 ± 43.2 | 0.002 |
| RWT | 0.43 ± 0.09 | 0.42 ± 0.09 | -0.01 ± 0.10 | 0.007 | 0.54 ± 0.10 | 0.51 ± 0.09 | -0.04 ± 0.10 | 0.197 |
| LAVI (mL/m<sup>2</sup>) | 39.6 ± 21.6 | 39.1 ± 17.9 | -0.5 ± 15.9 | 0.180 | 52.0 ± 15.2 | 46.5 ± 16.1 | -6.8 ± 14.8 | 0.123 |
| Mitral E/e' ratio | 12.8 ± 6.5 | 12.6 ± 8.3 | -0.2 ± 7.6 | 0.305 | 16.2 ± 8.1 | 13.0 ± 7.3 | -3.3 ± 6.0 | 0.064 |
| LV-GLS (%) | -14.7 ± 3.6 | -15.5 ± 3.2 | -0.8 ± 3.1 | <0.001 | -11.3 ± 3.0 | -15.2 ± 2.9 | -3.8 ± 3.6 | 0.002 |
| TR Vmax (m/s) | 2.38 ± 0.49 | 2.39 ± 0.61 | 0.01 ± 0.66 | 0.462 | 2.50 ± 0.46 | 2.53 ± 0.54 | 0.06 ± 0.47 | 0.673 |
| RVSP (mmHg) | 30.1 ± 9.4 | 30.0 ± 8.9 | 0.0 ± 8.5 | 0.979 | 32.4 ± 11.8 | 32.1 ± 11.9 | 0.4 ± 13.8 | 0.919 |
| **LV geometry** |  |  |  |  |  |  |  |  |
| Normal | 535 (34.6%) | 573 (37.1%) |  |  | 1 (7.1%) | 1 (7.1%) |  |  |
| Concentric remodeling | 326 (21.1%) | 307 (19.9%) |  |  | 2 (14.3%) | 6 (42.9%) |  |  |
| Concentric hypertrophy | 355 (23.0%) | 324 (21.0%) |  |  | 10 (71.4%) | 5 (35.7%) |  |  |
| Eccentric hypertrophy | 329 (21.3%) | 341 (22.1%) |  |  | 1 (7.1%) | 2 (14.3%) |  |  |
| **LV radius** |  |  |  |  |  |  |  |  |
| Basal (mm) | 53.6 ± 7.2 | 53.1 ± 6.6 | -0.5 ± 6.6 | 0.002 | 60.5 ± 7.9 | 55.8 ± 6.6 | -4.7 ± 5.2 | 0.005 |
| Mid (mm) | 57.1 ± 7.9 | 56.5 ± 7.3 | -0.6 ± 7.3 | 0.002 | 62.8 ± 7.3 | 58.1 ± 5.7 | -4.7 ± 5.1 | 0.004 |
| Apex (mm) | 38.0 ± 5.5 | 37.6 ± 5.1 | -0.3 ± 5.3 | 0.021 | 41.9 ± 5.0 | 39.4 ± 3.7 | -2.4 ± 3.8 | 0.031 |

| LV wall thickness |  |  |  |  |  |  |  |  |
| --- | --- | --- | --- | --- | --- | --- | --- | --- |
| Basal (mm) | 6.3 ± 1.0 | 6.2 ± 0.9 | -0.2 ± 1.1 | <0.001 | 8.0 ± 0.7 | 6.9 ± 0.9 | -1.1 ± 0.8 | <0.001 |
| Mid (mm) | 7.8 ± 1.2 | 7.7 ± 1.1 | -0.2 ± 1.1 | <0.001 | 9.8 ± 1.5 | 9.0 ± 0.9 | -0.8 ± 1.3 | 0.037 |
| Apex (mm) | 5.3 ± 1.0 | 5.2 ± 0.9 | -0.1 ± 0.9 | <0.001 | 6.0 ± 0.8 | 5.8 ± 1.1 | -0.2 ± 1.0 | 0.556 |
| Segmental longitudinal strain |  |  |  |  |  |  |  |  |
| Basal (%) | -14.1 ± 3.9 | -14.6 ± 3.8 | -0.6 ± 4.0 | <0.001 | -8.3 ± 2.1 | -13.4 ± 3.5 | -5.2 ± 3.4 | <0.001 |
| Mid (%) | -14.0 ± 3.9 | -14.9 ± 3.7 | -0.9 ± 3.9 | <0.001 | -7.5 ± 3.1 | -14.1 ± 2.7 | -6.6 ± 3.0 | <0.001 |
| Apex (%) | -16.3 ± 4.6 | -17.2 ± 4.1 | -0.9 ± 4.3 | <0.001 | -17.2 ± 4.1 | -19.4 ± 3.1 | -2.2 ± 4.3 | 0.078 |
| RRSR | 0.59 ± 0.14 | 0.59 ± 0.13 | 0.00 ± 0.14 | 0.622 | 1.11 ± 0.13 | 0.72 ± 0.10 | -0.39 ± 0.13 | <0.001 |
| LV segmental wall stress (σ) |  |  |  |  |  |  |  |  |
| Basal (mmHg) | 472.5 ± 98.5 | 423.8 ± 87.7 | -48.6 ± 108.9 | <0.001 | 542.9 ± 148.8 | 424.9 ± 61.7 | -118.0 ± 139.9 | 0.008 |
| Mid (mmHg) | 407.2 ± 87.0 | 362.9 ± 73.0 | -44.3 ± 89.4 | <0.001 | 463.8 ± 117.3 | 342.4 ± 67.0 | -121.4 ± 104.0 | <0.001 |
| Apex (mmHg) | 404.2 ± 101.9 | 359.2 ± 83.5 | -45.0 ± 104.2 | <0.001 | 505.3 ± 124.2 | 371.3 ± 107.2 | -134.0 ± 104.3 | <0.001 |

**Table 4.**
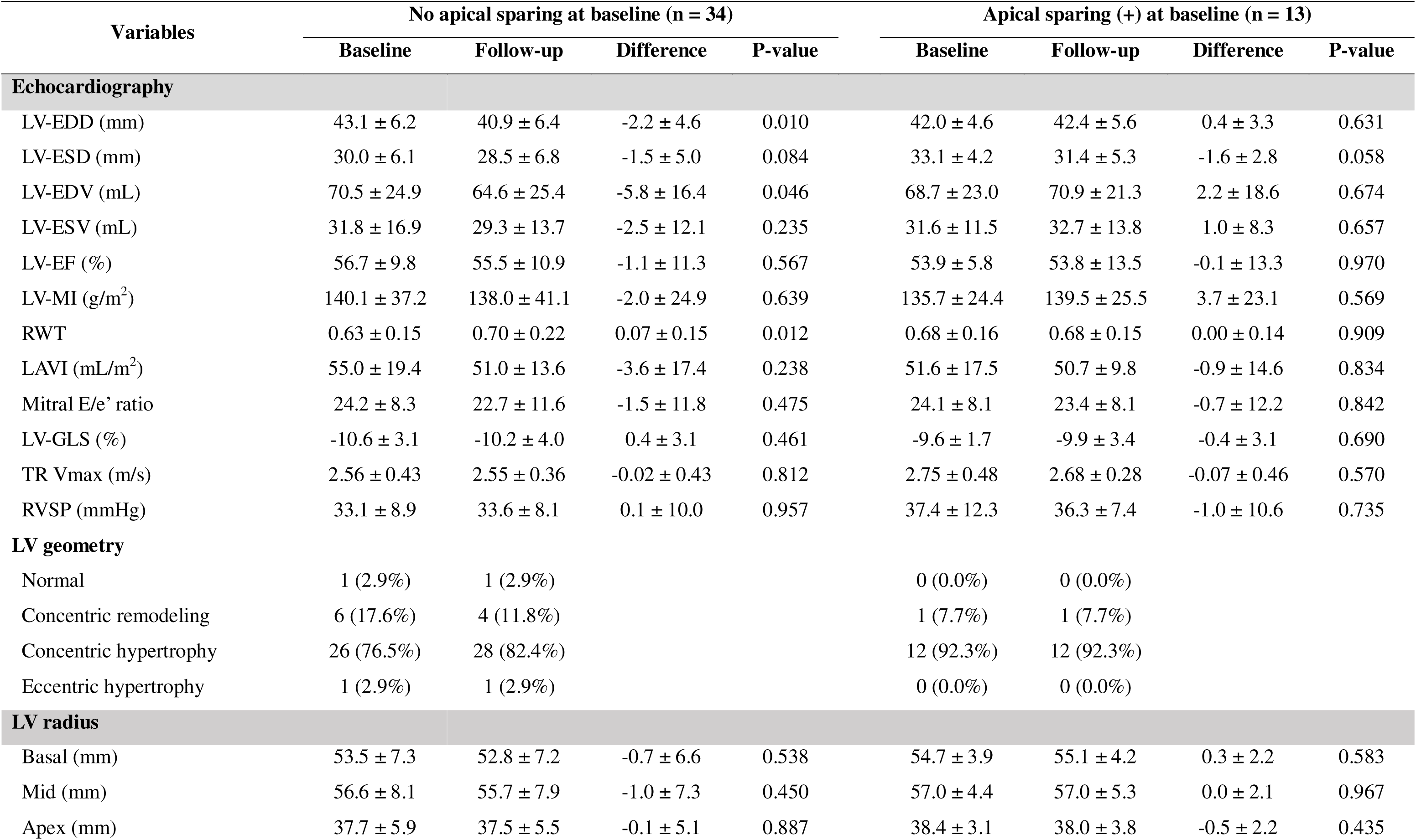
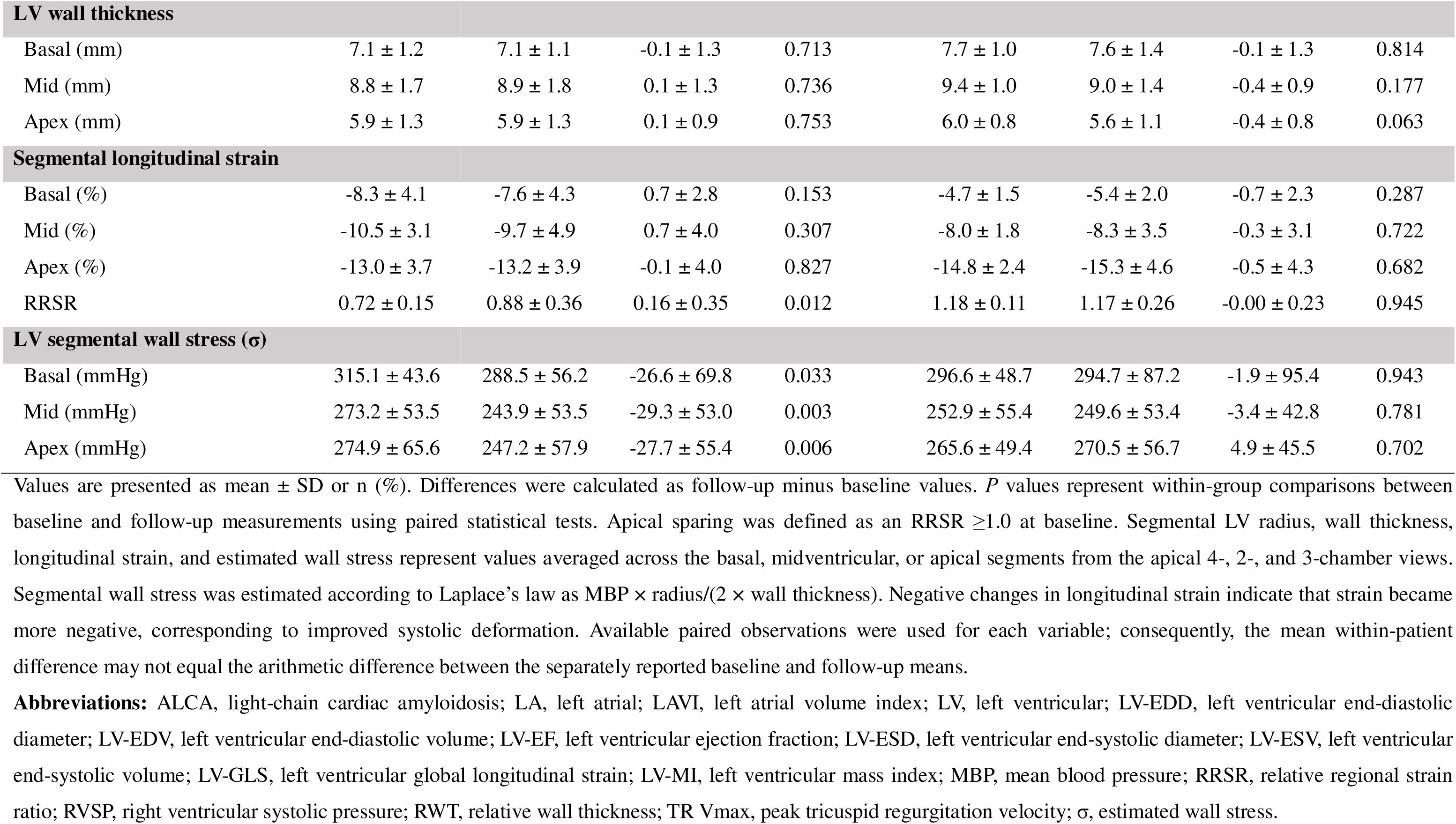
Temporal changes in echocardiographic parameters according to apical sparing pattern in patients with ALCA. Values are presented as mean ± SD or n (%). Differences were calculated as follow-up minus baseline values. *P* values represent within-group comparisons between baseline and follow-up measurements using paired statistical tests. Apical sparing was defined as an RRSR ≥1.0 at baseline. Segmental LV radius, wall thickness, longitudinal strain, and estimated wall stress represent values averaged across the basal, midventricular, or apical segments from the apical 4-, 2-, and 3-chamber views. Segmental wall stress was estimated according to Laplace’s law as MBP × radius/(2 × wall thickness). Negative changes in longitudinal strain indicate that strain became more negative, corresponding to improved systolic deformation. Available paired observations were used for each variable; consequently, the mean within-patient difference may not equal the arithmetic difference between the separately reported baseline and follow-up means. **Abbreviations:** ALCA, light-chain cardiac amyloidosis; LA, left atrial; LAVI, left atrial volume index; LV, left ventricular; LV-EDD, left ventricular end-diastolic diameter; LV-EDV, left ventricular end-diastolic volume; LV-EF, left ventricular ejection fraction; LV-ESD, left ventricular end-systolic diameter; LV-ESV, left ventricular end-systolic volume; LV-GLS, left ventricular global longitudinal strain; LV-MI, left ventricular mass index; MBP, mean blood pressure; RRSR, relative regional strain ratio; RVSP, right ventricular systolic pressure; RWT, relative wall thickness; TR Vmax, peak tricuspid regurgitation velocity; σ, estimated wall stress.

Temporal changes in the relationship between estimated wall stress and segmental LS are shown in **Figure 3**. In HHD, the positive associations observed at baseline were attenuated at follow-up at all LV levels (**Figure 3A**). In ALCA, higher wall stress tended to be associated with more preserved LS, and these relationships remained largely unchanged after treatment (**Figure 3B**).

**Figure 3.**
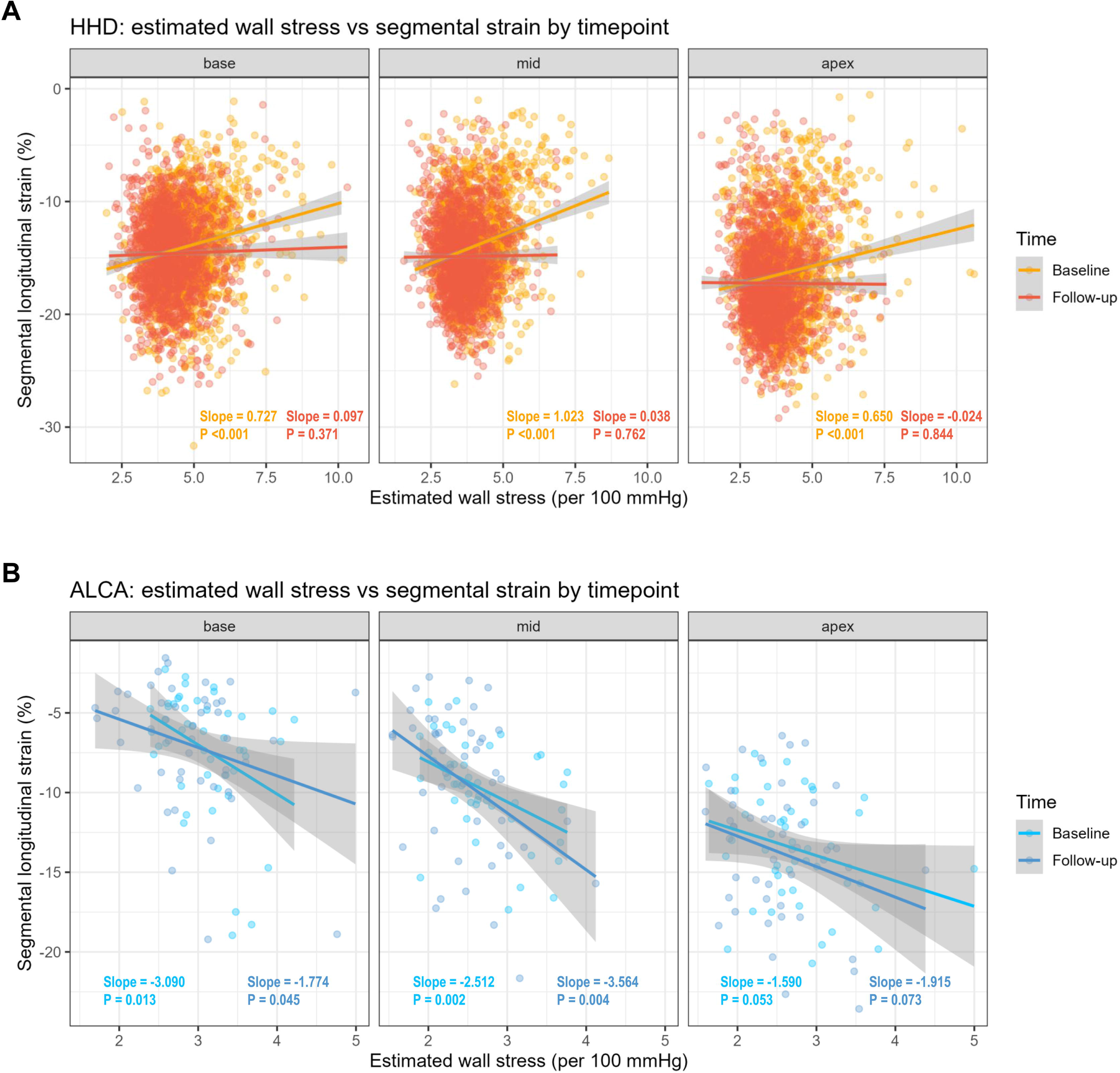
Associations of LV wall stress with segmental LS over time. Associations are shown at baseline and follow-up in (A) HHD and (B) ALCA. Abbreviations as in Figure 1.

### Predictors of Change in RRSR

Associations between changes in wall stress and segmental LS are shown in **Figure 4**. In HHD, changes in wall stress were associated with changes in LS at the midventricular and apical levels, but not at the basal level (basal slope −0.091, P=0.329; midventricular slope−0.303, P=0.006; apical slope −0.272, P=0.009) (**Figure 4A**). The association was more pronounced in patients with baseline apical sparing (**Figure 4B**). No significant associations were observed in ALCA (**Figures 4C**).

**Figure 4.**
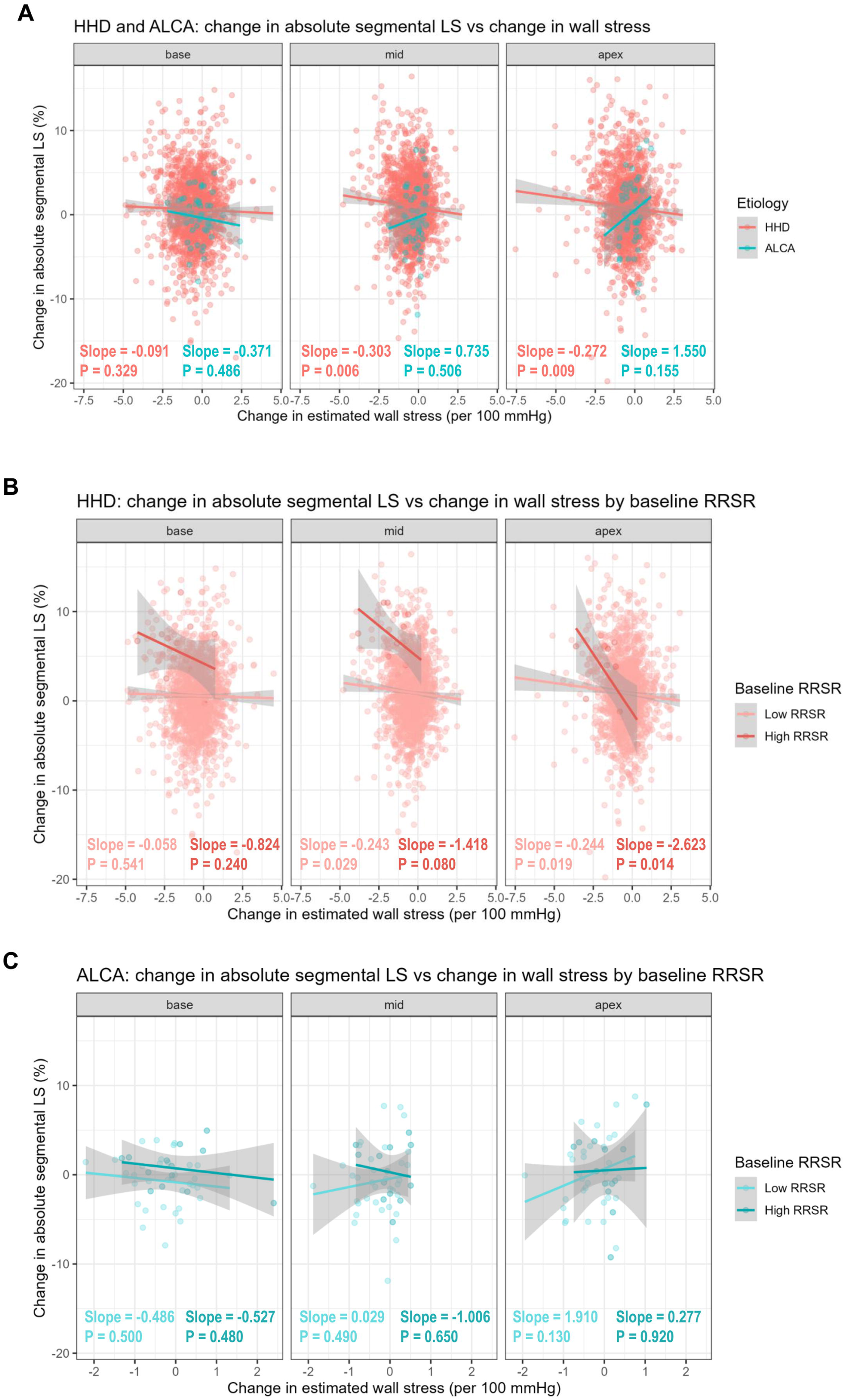
Associations of changes in wall stress with changes in segmental LS. Changes in wall stress and absolute segmental LS at the basal, midventricular, and apical levels are shown (A) by etiology, and according to baseline RRSR in (B) HHD and (C) ALCA. Abbreviations as in Figure 1.

Predictors of ΔRRSR are shown in **Table 5**. In HHD, diabetes mellitus (β=1.631; 95% CI: 0.063–3.200; P=0.042), an increase in MBP (β=1.569 per 10 mmHg; 95% CI: 0.997–2.141; P<0.001), and an increase in LVMI (β=0.367 per 10 g/m²; 95% CI: 0.150–0.583; P=0.001) were independently associated with increased RRSR. In ALCA, favorable hematologic response was the only independent determinant of RRSR reduction (β=−29.338; 95% CI: −46.683 to −11.993; P=0.001). Among patients with baseline apical sparing, ΔRRSR was associated with change in basal wall stress in HHD (β=0.267; 95% CI: 0.023–0.511; P=0.036) and with achievement of complete response or very good partial response in ALCA (β=−34.135; 95% CI: −58.948 to −9.321; P=0.012).

**Table 5.** Predictors of change in relative regional strain ratio in HHD and ALCA. Results for the entire HHD and ALCA populations were derived from multivariable linear regression analyses using parsimonious, clinically prespecified models. Because of the small sample sizes, analyses restricted to patients with apical sparing at baseline were performed using univariable linear regression and should be considered exploratory. The dependent variable was ΔRRSR × 100, calculated as (follow-up RRSR − baseline RRSR) × 100. Therefore, β coefficients represent the change in RRSR multiplied by 100. Positive β coefficients indicate an increase in RRSR during follow-up, whereas negative β coefficients indicate a reduction in RRSR. Changes in continuous variables were calculated as follow-up minus baseline values. Variables with substantial conceptual or mathematical overlap were not included simultaneously in the multivariable models. Apical sparing was defined as an RRSR ≥1.0 at baseline. Favorable hematologic response in ALCA was defined as CR or VGPR. Abbreviations: ALCA, light-chain cardiac amyloidosis; CI, confidence interval; CR, complete response; HHD, hypertensive heart disease; LVMI, left ventricular mass index; MBP, mean blood pressure; RRSR, relative regional strain ratio; VGPR, very good partial response; β, regression coefficient; Δ, change calculated as follow-up minus baseline.

| **Entire HHD population**<br>**(n = 1,559)** | **$\beta$** | **95% CI** | **P-value** | **Entire ALCA population**<br>**(n = 47)** | **$\beta$** | **95% CI** | **P-value** |
| --- | --- | --- | --- | --- | --- | --- | --- |
| Diabetes mellitus | 1.631 | 0.063 – 3.200 | 0.042 | CR or VGPR | -29.338 | -46.683 – -11.993 | 0.001 |
| ΔMBP (follow-up – baseline; per +10 mmHg) | 1.569 | 0.997 – 2.141 | <0.001 | | | | |
| ΔLVMI (follow-up – baseline; per +10 g/m<sup>2</sup>) | 0.367 | 0.150 – 0.583 | 0.001 | | | | |
| **HHD with apical sparing**<br>**at baseline**<br>**(n = 14)** | **$\beta$** | **95% CI** | **P-value** | **ALCA with apical sparing**<br>**at baseline**<br>**(n = 13)** | **$\beta$** | **95% CI** | **P-value** |
| ΔWall stress at basal segments (per +1 mmHg) | 0.267 | 0.023 – 0.511 | 0.036 | CR or VGPR | -34.135 | -58.948 – -9.321 | 0.012 |

Results for the entire HHD and ALCA populations were derived from multivariable linear regression analyses using parsimonious, clinically prespecified models. Because of the small sample sizes, analyses restricted to patients with apical sparing at baseline were performed using univariable linear regression and should be considered exploratory. The dependent variable was ΔRRSR × 100, calculated as (follow-up RRSR – baseline RRSR) × 100. Therefore, $\beta$ coefficients represent the change in RRSR multiplied by 100. Positive $\beta$ coefficients indicate an increase in RRSR during follow-up, whereas negative $\beta$ coefficients indicate a reduction in RRSR. Changes in continuous variables were calculated as follow-up minus baseline values. Variables with substantial conceptual or mathematical overlap were not included simultaneously in the multivariable models. Apical sparing was defined as an RRSR ≥ 1.0 at baseline. Favorable hematologic response in ALCA was defined as CR or VGPR.
Abbreviations: ALCA, light-chain cardiac amyloidosis; CI, confidence interval; CR, complete response; HHD, hypertensive heart disease; LVMI, left ventricular mass index; MBP, mean blood pressure; RRSR, relative regional strain ratio; VGPR, very good partial response; $\beta$ , regression coefficient; $\Delta$ , change calculated as follow-up minus baseline.

## Discussion

Using AI-assisted measurements of LV radius, wall thickness, and longitudinal strain, we estimated segmental LV wall stress in HHD and ALCA, with normal subjects as controls. The principal findings were as follows. First, apical sparing was observed in a small subset of patients with HHD, whereas it was substantially more frequent in ALCA and absent in normal controls, although prevalence decreased as the RRSR cutoff increased. Second, apical sparing in HHD was associated with pressure-overload features, whereas revised Mayo stage was its only independent determinant in ALCA. Third, in HHD, RRSR and apical-sparing prevalence decreased after antihypertensive treatment, particularly among patients with high baseline RRSR, with concomitant reductions in wall stress and improvement in basal and midventricular strain. Fourth, determinants of RRSR change differed by etiology, with changes in mean BP and wall stress associated with RRSR change in HHD and favorable hematologic response associated with RRSR reduction in ALCA. These findings suggest that apical sparing may represent a reversible, load-sensitive deformation pattern in HHD, which can be explained by wall stress according to Laplace law, and a disease-severity–related strain phenotype in ALCA.

### Apical Sparing Beyond Cardiac Amyloidosis

Apical sparing is widely recognized as an echocardiographic clue for cardiac amyloidosis, but it is not pathognomonic and has also been reported in severe aortic stenosis, chronic kidney disease, and other nonamyloid conditions.(1,5–7,26) Consistent with these reports, apical sparing was present in approximately 1% of patients with HHD using the conventional RRSR cutoff of 1.0.

The prevalence of apical sparing in our ALCA cohort was lower than in previous studies.(1,5,26) Earlier detection of cardiac involvement is one possible explanation, because both RRSR and apical-sparing prevalence increased during follow-up. However, this interpretation is tentative because temporal changes may also reflect disease progression, treatment, and hematologic response. Nonetheless, it should be noted that the prevalence of apical sparing in ALCA varied according to the RRSR threshold used, which was also reported by a previous study by Cotella J et al.(5) Similarly, although only 0.9% of patients with HHD met the conventional cutoff of RRSR ≥1.0, progressively larger proportions met lower thresholds of 0.7, 0.8, or 0.9. Thus, apical sparing is better viewed as a continuous deformation phenotype than a strictly binary finding. Lower thresholds may detect milder relative apical preservation but reduce specificity. Accordingly, both the absolute RRSR and the cutoff applied should be considered when comparing studies or interpreting apical sparing clinically.

These findings emphasize that apical sparing should be interpreted as a deformation phenotype rather than a disease-specific diagnosis. In patients with suspected cardiac amyloidosis, apical sparing remains an important red-flag sign requiring further evaluation, but its interpretation should incorporate clinical context, BP status, LV geometry, electrocardiographic findings, biomarkers, and multimodality imaging. As shown in the present study, apical sparing pattern can be observed in patients with HHD, in which severe pressure overload and elevated basal wall stress appeared to contribute to the regional strain pattern, whereas the apical sparing pattern in ALCA was associated with the stage of amyloidosis rather than BP. Thus, severe hypertension may produce a regional strain pattern that mimics the apical sparing commonly associated with cardiac amyloidosis.

The longitudinal findings further support this distinction. In HHD, both RRSR and the prevalence of apical sparing decreased from baseline to follow-up, and the reduction in RRSR was most evident among patients with higher baseline values. In contrast, both RRSR and apical-sparing prevalence increased in the overall ALCA population. Thus, temporal changes in apical sparing may provide mechanistic information beyond a single cross-sectional measurement.

### Segmental Wall Stress as a Mechanistic Framework in HHD

Laplace’s law, which implicates that wall stress increases with intracavitary pressure and radius and decreases with wall thickness, provides a plausible explanation for apical sparing in HHD.(9,27) The larger basal and midventricular radius may expose these regions to greater pressure-related stress than the apex, causing greater impairment of basal and midventricular strain and relative apical preservation.

Our findings support this framework. HHD showed the highest estimated wall stress across LV levels. Among patients with baseline apical sparing, basal and midventricular strain were markedly impaired, whereas apical strain was relatively preserved. After antihypertensive treatment, RRSR and apical-sparing prevalence decreased, basal and midventricular strain improved, and LV dimensions, LVMI, radius, and wall stress declined. These changes are consistent with prior evidence linking BP reduction to LV reverse remodeling and LVH regression.(12,13) Higher mean BP independently predicted baseline apical sparing, while changes in mean BP and segmental wall stress were associated with RRSR change. The reduction in RRSR was driven mainly by recovery of basal and midventricular strain rather than by substantial apical change and was greatest in patients with higher baseline RRSR. Together, these findings suggest that apical sparing in HHD is a dynamic, load-sensitive, and potentially reversible phenotype compatible with Laplace’s law.

### Distinct Determinants of Apical Sparing in ALCA

The mechanism of apical sparing in ALCA appeared different. Despite greater wall thickness, smaller LV cavities, lower BP, and lower estimated wall stress than HHD, patients with ALCA had more impaired global longitudinal strain and more frequent apical sparing. Prior PET and CMR studies suggest that amyloid-related apical sparing reflects regional differences in amyloid burden or total amyloid mass rather than pressure-derived wall stress.(3) In the present study, revised Mayo stage was the only independent determinant of baseline apical sparing, consistent with the importance of cardiac biomarkers and free light-chain burden in ALCA.(18,19,28) Moreover, favorable hematologic response was the only determinant of RRSR reduction during follow-up.(19) Thus, the overall increase in RRSR may have been driven by patients with persistent or progressive disease, whereas those achieving a favorable response improved. These findings suggest that apical sparing in ALCA reflects disease severity and treatment response rather than loading conditions.

### Clinical Implications

The present findings have several clinical implications. First, apical sparing should not be considered specific to cardiac amyloidosis, although it remains an important clue to amyloid cardiomyopathy. Second, the apparent prevalence and clinical meaning of apical sparing depend on the RRSR cutoff: the conventional threshold of 1.0 identifies a pronounced but uncommon phenotype in HHD, whereas lower thresholds capture a broader and less specific spectrum. Third, BP and LV geometry should therefore be considered when interpreting apical sparing. In patients with high BP, increased LV radius, and elevated wall stress, the pattern may reflect regional load-dependent dysfunction rather than infiltration. Fourth, serial strain assessment may provide additional insight: resolution after BP reduction supports a load-sensitive mechanism consistent with Laplace’s law. Finally, segmental wall-stress analysis may therefore provide a physiologic framework for interpreting regional strain abnormalities in pressure overload and infiltrative disease. Although absolute RRSR values may vary by vendor and analytic method, within-patient longitudinal changes measured consistently may be more informative than a single cross-sectional cutoff.

### Limitations

Several limitations should be acknowledged. First, its retrospective observational design precludes causal inference. Second, apical sparing was rare in HHD, limiting the stability of multivariable and subgroup analyses. Third, wall stress was estimated using a simplified Laplace-based model with mean BP as a surrogate for LV pressure and two-dimensional measures of LV geometry. Invasive pressure, transmural pressure, three-dimensional geometry, myocardial material properties, and fiber orientation were unavailable. Finally, tissue-level amyloid burden was not available in all patients with ALCA, precluding direct correlation with RRSR and estimated wall stress.

## Conclusions

Apical sparing occurred in a small subset of patients with HHD and regressed after antihypertensive treatment. In HHD, it was associated with pressure-overload mechanics, and RRSR reduction accompanied decreased segmental wall stress and improved basal and midventricular strain, supporting a Laplace-based mechanism. In ALCA, apical sparing was associated with revised Mayo stage, and RRSR reduction was linked to favorable hematologic response. Thus, a similar regional strain phenotype may represent a reversible, load-sensitive pattern in HHD and a disease-severity–related phenotype in ALCA.

## Abbreviations

ALCA: light-chain cardiac amyloidosis
BP: blood pressure
CI: confidence interval
DBP: diastolic blood pressure
eGFR: estimated glomerular filtration rate
HHD: hypertensive heart disease
LA: left atrial
LAVI: left atrial volume index
LS: longitudinal strain
LV: left ventricular
LVEF: left ventricular ejection fraction
LVH: left ventricular hypertrophy
LVMI: left ventricular mass index
MBP: mean blood pressure
OR: odds ratio
RRSR: relative regional strain ratio
RWT: relative wall thickness
SBP: systolic blood pressure
SD: standard deviation

## Perspectives

### Competency in Medical Knowledge

Apical sparing of left ventricular longitudinal strain is not specific to cardiac amyloidosis. In hypertensive heart disease, it may represent a reversible, load-sensitive pattern of regional dysfunction related to elevated wall stress. Accordingly, blood pressure, left ventricular geometry, and multimodality imaging findings should be integrated when interpreting this strain pattern.

### Translational Outlook

Prospective studies are needed to determine whether serial assessment of segmental wall stress and longitudinal strain improves discrimination between pressure-overload–related apical sparing and infiltrative myocardial disease, and whether artificial intelligence–based analysis can facilitate individualized evaluation of left ventricular hypertrophy.

## Data Availability

The datasets used and analyzed during the current study are available from the corresponding author on reasonable request.

## Acknowledgments

We thank Lia Ju, a registered diagnostic cardiac sonographer (RDCS), for her dedication and valuable support in the conduct of this study.

## Sources of Funding

This research did not receive any specific grant from funding agencies in the public, commercial, or not-for-profit sectors.

## Conflict of Interest

Y.J., J.J., S.A.L., and Y.E.Y are currently affiliated with Ontact Health Co., Ltd. Y.J. and S.A.L. are co-inventors on patents related to this work filed by Ontact Health Co., Ltd.: Korean patent application KR10-2024-0115280, “Method for providing information on strain quantification and device for providing information on strain quantification using the same,” for which a decision to grant has been issued but registration has not yet been completed; and U.S. patent application US19/229913, “METHOD AND DEVICE FOR PROVIDING INFORMATION ON STRAIN QUANTIFICATION,” currently pending. Y.E.Y holds equity in Ontact Health Co., Ltd. The other authors have no conflicts of interest to declare.

---

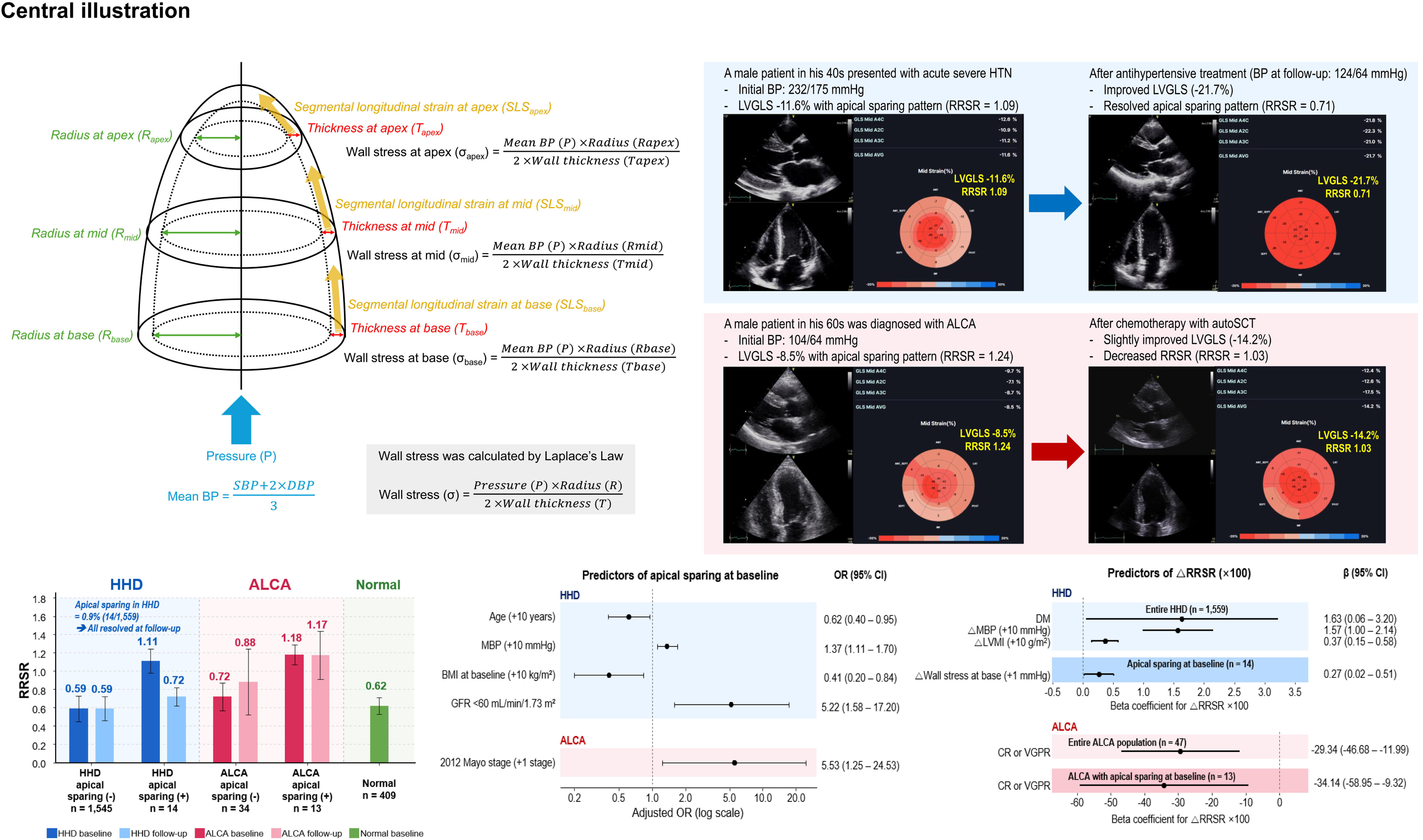

## Notes

### Author Declarations

Institutional Review Board of the Clinical Research Institute at Seoul National University Bundang Hospital (No. B-2206-762-102 / No. B-2208-773-108) and the Institutional Review Board at Chung-Ang University Hospital (No. 2205-014-19419) gave ethical approval for this work, and waived the requirement for informed consent, given the retrospective nature of the study and minimal expected risk to the participants.

