## Supplementary figure and table for "Transient Apical Sparing in Hypertensive Heart Disease Explained by Laplace’s Law"

**Short Title:** Apical sparing in HHD explained by Laplace's law

In-Chang Hwang[a,b*], Hyue Mee Kim[c*], Yeonggul Jang[d], Minjung Bak[a,b], Jiesuck Park[a,b], Jaeik Jeon[d], Seung-Ah Lee[d], Hong-Mi Choi[a,b], Yeonyee E. Yoon[a,b,d], Goo-Yeong Cho[a,b]

[a] Department of Cardiology, Cardiovascular Center, Seoul National University Bundang Hospital, Seongnam, Gyeonggi;

[b] Department of Internal Medicine, Seoul National University College of Medicine, Seoul;

[c] Division of Cardiology, Department of Internal Medicine, Chung-Ang University Hospital, Seoul, South Korea

[d] Ontact Health Co, Ltd., Seoul, Republic of Korea

[*] These two authors equally contributed to this work as co-corresponding authors.

### Supplementary Table S1. Detailed clinical profiles of ALCA group

| Variables | ALCA (n=47) | ALCA without apical sparing (n=34) | ALCA with apical sparing (n=13) | P |
|---|---|---|---|---|
| **Clinical Factors** | | | | |
| Age (years) | 71.4 ± 9.2 | 72.4 ± 9.4 | 68.6 ± 8.3 | 0.212 |
| Male sex | 29 (61.7%) | 19 (55.9%) | 10 (76.9%) | 0.315 |
| Hypertension | 23 (48.9%) | 15 (44.1%) | 8 (61.5% | 0.285 |
| Diabetes mellitus | 10 (21.3%) | 8 (23.5%) | 2 (15.4%) | 0.703 |
| Dyslipidemia | 16 (34.0%) | 11 (32.4%) | 5 (38.5%) | 0.693 |
| Chronic kidney disease | 18 (38.3%) | 14 (41.2%) | 4 (30.8%) | 0.739 |
| Coronary artery disease | 1 (2.1%) | 1 (2.9%) | 0 (0.0%) | 0.723 |
| Atrial fibrillation | 8 (17.0%) | 3 (8.8%) | 5 (38.5%) | 0.028 |
| Stroke | 5 (10.6%) | 2 (5.9%) | 3 (23.1%) | 0.121 |
| **Anthropometric Factors** | | | | |
| Body mass index (kg/m$^2$) | 24.1 ± 2.5 | 23.6 ± 2.5 | 23.3 ± 2.4 | 0.717 |
| SBP at baseline (mmHg) | 113.7 ± 20.6 | 114.3 ± 19.3 | 112.3 ± 24.5 | 0.774 |
| DBP at baseline (mmHg) | 68.4 ± 9.8 | 68.4 ± 8.5 | 68.5 ± 13.0 | 0.962 |
| MBP at baseline (mmHg) | 83.5 ± 12.4 | 83.7 ± 10.7 | 83.1 ± 16.4 | 0.891 |
| **Baseline echocardiography** | | | | |
| IVSd (mm) | 14.3 ± 2.6 | 14.2 ± 2.8 | 14.6 ± 2.1 | 0.634 |
| LV-EDD (mm) | 42.8 ± 5.8 | 43.1 ± 6.2 | 42.0 ± 4.6 | 0.572 |
| LV-ESD (mm) | 30.8 ± 5.8 | 30.0 ± 6.1 | 33.1 ± 4.2 | 0.101 |
| LVPWd (mm) | 13.5 ± 2.2 | 13.3 ± 2.3 | 13.9 ± 2.1 | 0.363 |
| LV-MI (g/m$^2$) | 138.9 ± 33.9 | 140.1 ± 37.2 | 135.7 ± 24.4 | 0.698 |
| RWT | 0.64 ± 0.15 | 0.63 ± 0.15 | 0.68 ± 0.16 | 0.336 |
| LV-EDV (mL) | 70.0 ± 24.2 | 70.5 ± 24.9 | 68.7 ± 23.0 | 0.828 |
| LV-ESV (mL) | 31.7 ± 15.5 | 31.8 ± 16.9 | 31.6 ± 11.5 | 0.977 |
| LV-EF (%) | 55.9 ± 8.9 | 56.7 ± 9.8 | 53.9 ± 5.8 | 0.347 |
| LA diameter (mm) | 42.6 ± 5.4 | 41.7 ± 5.7 | 44.8 ± 3.5 | 0.078 |
| LAVI (mL/m$^2$) | 54.1 ± 18.8 | 55.0 ± 19.4 | 51.6 ± 17.5 | 0.585 |
| Mitral E/e' ratio | 24.2 ± 8.1 | 24.2 ± 8.3 | 24.1 ± 8.1 | 0.981 |
| LV-GLS (%) | -10.3 ± 2.8 | -10.6 ± 3.1 | -9.6 ± 1.7 | 0.158 |
| TR Vmax (m/s) | 2.6 ± 0.4 | 2.6 ± 0.4 | 2.8 ± 0.5 | 0.180 |
| RVSP (mmHg) | 34.4 ± 10.1 | 33.1 ± 8.9 | 37.4 ± 12.3 | 0.206 |
| **LV geometry at baseline** | | | | 0.641 |
| Normal | 1 (2.1%) | 1 (2.9%) | 0 (0.0%) | |
| Concentric remodeling | 7 (14.9%) | 6 (17.6%) | 1 (7.7%) | |
| Concentric hypertrophy | 38 (80.9%) | 26 (76.5%) | 12 (92.3%) | |
| Eccentric hypertrophy | 1 (2.1%) | 1 (2.9%) | 0 (0.0%) | |
| **Antihypertensive medication** | | | | |
| RAS blockers | 14 (29.8%) | 14 (29.8%) | 0 (0.0%) | <0.001 |
| Beta blockers | 10 (21.3%) | 10 (21.3%) | 0 (0.0%) | <0.001 |
| CCB | 10 (21.3%) | 10 (21.3%) | 0 (0.0%) | <0.001 |
| Thiazide | 1 (2.1%) | 1 (2.1%) | 0 (0.0%) | <0.001 |
| MRA | 19 (40.4%) | 19 (40.4%) | 0 (0.0%) | <0.001 |
| **Chemotherapy for ALCA** | | | | |
| **AutoSCT** | 10 (21.3%) | 6 (17.6%) | 4 (30.8%) | 0.325 |
| **Daratumumab-containing regimen** | 2 (4.3%) | 1 (2.9%) | 1 (7.7%) | 0.470 |
| - Dara-VTD (Daratumumab + Bortezomib + Thalidomide + Dexamethasone) | 1 (2.1%) | 1 (2.9%) | 0 (0.0%) | 0.532 |
| - DRd (Daratumumab + Lenalidomide + Dexamethasone) | 1 (2.1%) | 0 (0.0%) | 1 (7.7%) | 0.102 |
| **PI-based regimen without daratumumab** | 33 (70.2%) | 24 (70.6%) | 9 (69.2%) | 0.927 |
| - Vel-Cytox-D (Bortezomib + Cyclophsophamide + Dexamethasone) | 3 (6.4%) | 1 (2.9%) | 2 (15.4%) | 0.119 |
| - VTD (Bortezomib + Thalidomide + Dexamethasone) | 3 (6.4%) | 1 (2.9%) | 2 (15.4%) | 0.119 |
| - VRd (Bortezomib + Lenalidomide + Dexamethasone) | 3 (6.4%) | 2 (5.9%) | 1 (7.7%) | 0.820 |
| - IRd (Ixazomib + Lenalidomide + Dexamethasone) | 4 (8.5%) | 4 (11.8%) | 0 (0.0%) | 0.196 |
| - Vel-Dex (Bortezomib + Dexamethasone) | 3 (6.4%) | 2 (5.9%) | 1 (7.7%) | 0.820 |
| - VMP (Bortezomib + Melphalan + | 23 (48.9%) | 17 (50.0%) | 6 (46.2%) | 0.813 |

| | | | | |
|---|---|---|---|---|
| Prednisolone) | | | | |
| **Non-PI combination or monotherapy** | 16 (34.0%) | 12 (35.3%) | 4 (30.8%) | 0.770 |
| - Mel-Dex (Melphalan + Dexamethasone) | 7 (14.9%) | 6 (17.6%) | 1 (7.7%) | 0.391 |
| - Cytox-Dex (Cyclophosphamide + Dexamethasone) | 1 (2.1%) | 1 (2.9%) | 0 (0.0%) | 0.532 |
| - Rd (Lenalidomide + Dexamethasone) | 3 (6.4%) | 1 (2.9%) | 2 (15.4%) | 0.119 |
| - Rev (Lenalidomide monotherapy) | 4 (8.5%) | 3 (8.8%) | 1 (7.7%) | 0.901 |
| - Thal-Dex (Thalidomide + Dexamethasone) | 2 (4.3%) | 1 (2.9%) | 1 (7.7%) | 0.470 |
| - Thalidomide monotherapy | 2 (4.3%) | 1 (2.9%) | 1 (7.7%) | 0.470 |
| - Dexamethasone monotherapy | 1 (2.1%) | 1 (2.9%) | 0 (0.0%) | 0.532 |
| **Revised Mayo Stage (2012)** | | | | 0.104 |
| - Stage I | 0 (0.0%) | 0 (0.0%) | 0 (0.0%) | |
| - Stage II | 4 (8.5%) | 4 (11.8%) | 0 (0.0%) | |
| - Stage III | 18 (38.3%) | 15 (44.1%) | 3 (23.1%) | |
| - Stage IV | 25 (53.2%) | 15 (44.1%) | 10 (76.9%) | |
| **Biomarkers** | | | | |
| - Troponin I | 0.17 (0.09 – 0.29) | 0.16 (0.07 – 0.25) | 0.19 (0.15 – 0.28) | 0.577 |
| - Troponin T | 0.09 (0.05 – 0.19) | 0.12 (0.05 – 0.20) | 0.09 (0.05 – 0.14) | 0.436 |
| - NT-proBNP (pg/mL) | 7448.5 (2872.7 – 14596.7) | 5917.0 (2639.6 – 12042.0) | 9059.9 (3603.0 – 20427.0) | 0.270 |
| - Difference (kappa vs. lambda) | 350.0 (1325 – 666.2) | 293.8 (122.0 – 666.2) | 460.2 (294.7 – 594.0) | 0.501 |
| **Response** | | | | 0.474 |
| - Complete response (CR) | 11 (23.4%) | 6 (17.6%) | 5 (38.5%) | |
| - Very good partial response (VGPR) | 9 (19.1%) | 6 (17.6%) | 3 (23.1%) | |
| - No response | 27 (57.4%) | 22 (64.7%) | 5 (38.5%) | |

Values are presented as mean ± SD, median (interquartile range), or n (%), as appropriate. P values compare patients with ALCA without versus with apical sparing at baseline and were calculated using the Student t test or Mann–Whitney U test for continuous variables and the chi-square test or Fisher exact test for categorical variables, as appropriate. Apical sparing was defined as an RRSR ≥1.0. Chronic kidney disease was defined as an eGFR <60 mL/min/1.73 m² at baseline. MBP was calculated as (SBP + 2 × DBP)/3. LV geometry was classified according to LVMI and RWT. Chemotherapy categories and individual regimens were not mutually exclusive because some patients received more than 1 treatment regimen during follow-up. Hematologic response was classified according to established response criteria; no response included partial response, stable disease, and progressive disease.

Abbreviations: ALCA, light-chain cardiac amyloidosis; AutoSCT, autologous stem cell transplantation; CCB, calcium channel blocker; CR, complete response; DBP, diastolic blood pressure; Dara-VTD, daratumumab, bortezomib, thalidomide, and dexamethasone; DRd, daratumumab, lenalidomide, and dexamethasone; eGFR, estimated glomerular filtration rate; IRd, ixazomib, lenalidomide, and dexamethasone; IVSd, interventricular septal thickness at end-diastole; LA, left atrial; LAVI, left atrial volume index; LV, left ventricular; LV-EDD, left ventricular end-diastolic diameter; LV-EDV, left ventricular end-diastolic volume; LV-EF, left ventricular ejection fraction; LV-ESD, left ventricular end-systolic diameter; LV-ESV, left ventricular end-systolic volume; LV-GLS, left ventricular global longitudinal strain; LV-MI, left ventricular mass index; LVPWd, left ventricular posterior wall thickness at end-diastole; MBP, mean blood pressure; Mel-Dex, melphalan and dexamethasone; MRA, mineralocorticoid receptor antagonist; NT-proBNP, N-terminal pro–B-type natriuretic peptide; PI, proteasome inhibitor; RAS, renin–angiotensin system; Rd, lenalidomide and dexamethasone; RRSR, relative regional strain ratio; RVSP, right ventricular systolic pressure; RWT, relative wall thickness; SBP, systolic blood pressure; Thal-Dex, thalidomide and dexamethasone; TR Vmax, peak tricuspid regurgitation velocity; Vel-Cytox-D, bortezomib, cyclophosphamide, and dexamethasone; Vel-Dex, bortezomib and dexamethasone; VGPR, very good partial response; VMP, bortezomib, melphalan, and prednisolone; VRd, bortezomib, lenalidomide, and dexamethasone; VTD, bortezomib, thalidomide, and dexamethasone.

**Supplementary Figure S1. Segment-based assessment of LV wall thickness, radius, wall stress, and longitudinal strain across HHD, ALCA, and normal groups**

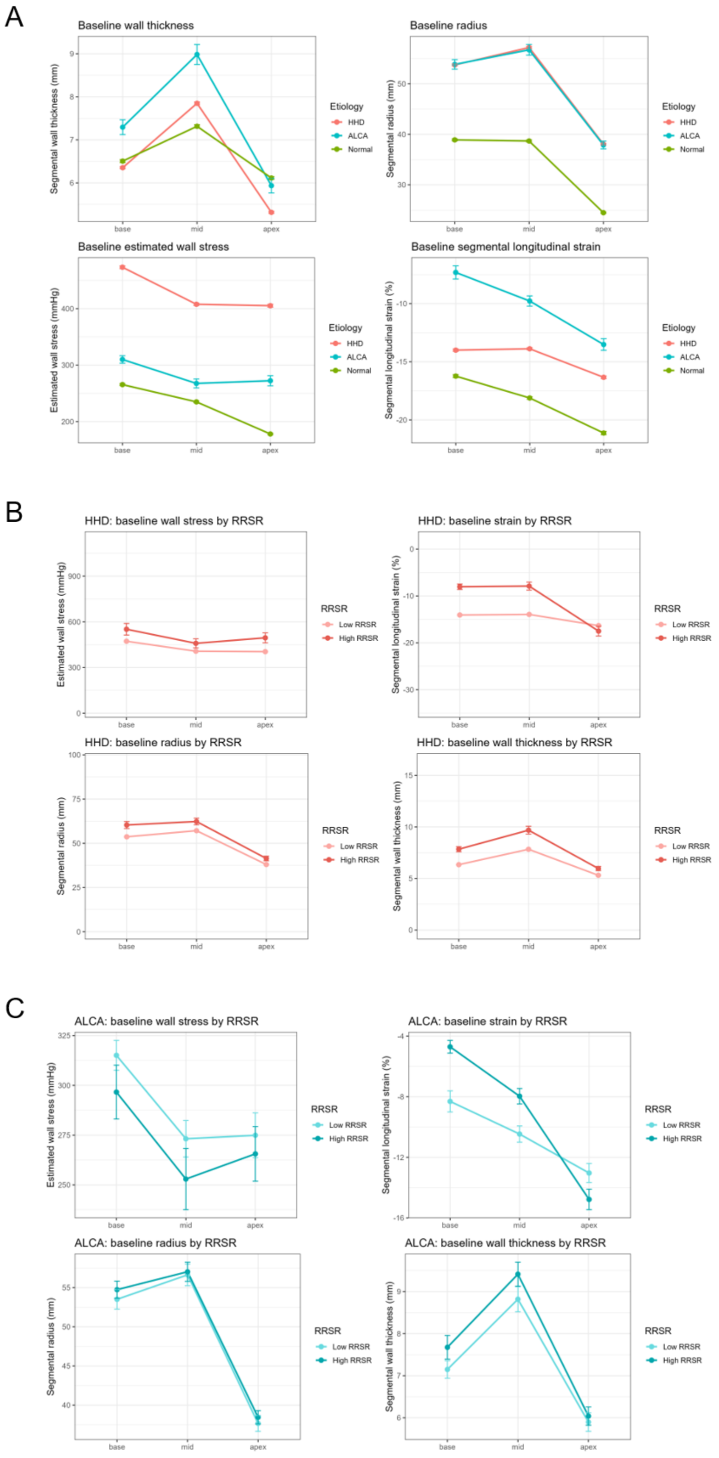


(A) Segment-based LV wall thickness, radius, wall stress, and longitudinal strain are compared among the HHD (red), ALCA (blue), and normal (green) groups. (B) Within the HHD group, segment-based measurements are compared according to the presence of apical sparing (RRSR <1.0, light red; RRSR ≥1.0, dark red). (C) Within the ALCA group, segment-based measurements are compared according to the presence of apical sparing (RRSR <1.0, light blue; RRSR ≥1.0, dark blue).
